# Adapting COVID-19 research infrastructure to capture influenza and respiratory syncytial virus alongside SARS-CoV-2 in UK healthcare workers winter 2022/23: Results of a pilot study in the SIREN cohort

**DOI:** 10.1101/2024.12.09.24318698

**Authors:** Sarah Foulkes, Katie Munro, Dominic Sparkes, Jonathan Broad, Naomi Platt, Anna Howells, Omolola Akinbami, Jameel Khawam, Palak Joshi, Sophie Russell, Chris Norman, Lesley Price, Diane Corrigan, Michelle Cole, Jean Timelin, Louise Forster, Katrina Slater, Conall H Watson, Nick Andrews, Andre Charlett, SIREN study group, Ana Atti, Jasmin Islam, Colin S Brown, Jonathan Turner, Susan Hopkins, Victoria Hall

**Affiliations:** United Kingdom Health Security Agency (UKHSA), 10 South Colonnade, Canary Wharf, London, United Kingdom, E14 4PU; Health and Care Research Wales, 5 15, Castlebridge, 19 Cowbridge Rd E, Cardiff CF11 9AB; Public Health Scotland, Meridian Court, 5 Cadogan Street, Glasgow, United Kingdom, G2 6QE; Health and Social Care in Northern Ireland (HSC), 12-22 Linenhall Street, Belfast, United Kingdom, BT2 8BS

## Abstract

**Introduction:** The combination of patient illness and staff absence driven by seasonal viruses culminates in annual “winter pressures” on UK healthcare systems and has been exacerbated by COVID-19. In winter 2022/23 we ran a pilot study aiming to introduce multiplex testing to determine the incidence and burden of SARS-CoV-2, influenza and respiratory syncytial virus (RSV) in our cohort of UK healthcare workers (HCWs).

**Methods:** The pilot study was conducted from 28/11/2022-31/03/2023 within the SIREN prospective cohort study. Participants completed fortnightly questionnaires, capturing symptoms and sick leave, and multiplex PCR testing for SARS-CoV-2, influenza and RSV, regardless of symptoms. PCR-positivity rates by virus were calculated over time, and viruses were compared by symptoms and severity. Self-reported symptoms and associated sick leave were described. Sick leave rates were compared by vaccination status and demographics.

**Results:** 5,863 participants were included, 84.6% female, 70.3% ≥45-years, and 33.4% were nurses. PCR-positivity peaked in early December for all three viruses (4.6 positives per 100 tests (95%CI 3.5, 5.7) SARS-CoV-2, 3.9 (95%CI 2.2, 5.6) influenza, 1.4 (95%CI 0.4, 2.4) RSV), declining to <0.3/100 tests after January for influenza/RSV, and around 2.5/100 tests for SARS-CoV-2. Over one-third of all infections were asymptomatic, and symptoms were similar for all viruses. 1,368 (23.3%) participants reported taking sick leave, median 4 days (range 1-59). Rates of sick leave were higher in participants with co-morbidities, working in clinical settings, and who had not been vaccinated (COVID-19 booster or seasonal influenza vaccine) versus those who had received neither vaccine (2.04 vs 1.41 sick days/100 days, adjusted Incidence Rate Ratio 1.47 (95%CI 1.38, 1.56).

**Conclusion:** This pilot demonstrated the use of multiplex testing allowed better understanding of the impact of seasonal respiratory viruses and respective vaccines on the HCW workforce. This highlights the important information on asymptomatic infection and persisting levels of SARS-CoV-2 infection.

## INTRODUCTION

UK healthcare systems experience significant challenges each winter, due to the impact of seasonal surges in respiratory viruses, including influenza and respiratory syncytial virus (RSV), which combined are often described as “winter pressures” (1, 2). Respiratory illness increases patient attendance but also causes significant staff absence, with respiratory illness the second most common cause of sick leave within the NHS (3). Reduction in staff sickness absence and preventing nosocomial transmission are the rationale for the annual NHS winter flu vaccine campaign for healthcare workers (HCW) and the prioritisation of HCW for COVID-19 vaccine boosters (4, 5).

As a result of the pandemic, COVID-19 has been well characterised in relation to key scientific and clinical questions including the role of antibodies as correlates of protection, rates of asymptomatic disease and real-world vaccine effectiveness (6–11). However, this is not the case for the other principle seasonal viruses, influenza and RSV, where the focus has been on symptomatic disease and in populations most at risk, including children and the elderly (12, 13). HCWs are an important population to study given the impact of respiratory illnesses on the workforce and their high exposure (14, 15), offering insights into the natural history, epidemiology and burden of influenza and RSV in working aged populations.

The SARS-CoV-2 Immunity and Reinfection Evaluation (SIREN) study is a prospective cohort study of HCWs across the UK, with participants completing regular SARS-CoV-2 PCR testing and antibody testing continuously since June 2020 (16). It was set up at the start of the pandemic to assess the risk of re-infection with SARS-CoV-2 and has continued to adapt to address key scientific questions.

During the COVID-19 pandemic, the introduction of non-pharmaceutical measures, including universal masking and social distancing, was found to reduce the rates of other respiratory viruses (4, 17). However, following the removal of these interventions, the winter of 2022/23 was the first opportunity to understand the impact of circulating seasonal viruses on the NHS workforce. Therefore, in winter 2022/23 the SIREN study piloted a “Winter Pressures sub-study” that included multiplex PCR testing for influenza and RSV alongside SARS-CoV-2 (18).

This paper outlines the results of the SIREN Winter Pressures Pilot study 2022/23, with the aim of a) describing the burden of influenza (A and B), RSV, and SARS-CoV-2, b) characterisation of SIREN HCW symptom profile, and c) understanding time off work due to being symptomatic in the context of HCW vaccination.

## METHODS

### Study design

The SIREN Winter Pressures sub-study is a prospective cohort study nested within the SIREN UK multicentre HCW cohort study (16, 18).

### Participants

Participants were recruited into the sub-study via two routes: 1) participants previously completing monoplex testing (for SARS-CoV-2 only) were informed, in writing, before the sub-study start date (28 November 2022) that testing would move to multiplex testing (SARS-CoV-2, Influenza and RSV), and were given the opportunity to withdraw from the study; 2) additional participants were re-recruited (from participants who were not currently testing) and consented directly into the winter pressures postal testing pathway, between 13 December 2022 and 24 January 2023 (18).

Participants undergoing PCR testing between 28 November 2022 and 31 March 2023 were included in the sub-study. Participants contributed different lengths of time to the analysis period, due to completing study follow-up time or withdrawing from the study.

### Data collection

Participants completed fortnightly questionnaires on symptoms (onset date, type and duration) and related time taken-off work due to reported symptoms, in addition to an enrolment survey at the start of the SIREN study, detailing the participant’s demographics, occupation, underlying medical conditions and household composition. Vaccination data (COVID-19 and seasonal influenza vaccination) was obtained both from the fortnightly questionnaire and linkage to national vaccination registries.

Participants completed swabs for either multiplex PCR (SARS-CoV-2, influenza and where available RSV) or monoplex PCR (SARS-CoV-2 only), fortnightly regardless of symptoms. PCR testing, symptoms and sociodemographic data were linked via a unique study ID.

### Variables

We defined participants who reported at least one of any of the following symptoms as being symptomatic: cough, fever, shortness of breath, sore throat, runny nose, headache, muscle aches, altered sense of smell or taste, fatigue, diarrhoea, nausea or vomiting, itchy red patches on fingers or toes, rash, swollen glands (19). Influenza-like illness (ILI) was defined as participants reporting fever and at least one of following: cough, sore throat, shortness of breath, headache, muscle ache or fatigue. For this analysis, COVID-19 vaccination refers to receiving the second booster dose of the COVID-19 vaccine, and influenza vaccination refers to receiving the 2022/23 seasonal influenza vaccine between 01 September 2022 and 31 March 2023. Sick leave was defined as any self-reported days off work due to being symptomatic.

### Outcomes

The co-primary outcomes were a) a PCR-positive test for either SARS-CoV-2, influenza or RSV; b) proportion of participants reporting ILI symptoms; and c) the number of days taken off work due to being symptomatic.

### Inclusion criteria

Participants who completed at least one fortnightly questionnaire and at least one PCR test during the analysis period were included.

### Statistical analysis

Participants’ sociodemographic and occupational characteristics were described.

Infection rates by pathogen were calculated over time. Samples were initially de-duplicated resulting in one test per fortnight per participant, prioritising a positive sample over a negative sample in the same fortnight. Samples which were positive for multiple viruses were excluded from the analysis. Further de-duplication by infection episode meant that for SARS-CoV-2, participants could only have one positive sample per 90 days. For influenza and RSV, participants could only have one positive sample per 30 days.

For the infection analysis, participants were considered vaccinated if they had received the seasonal vaccine dose at least 14 days before the first positive PCR sample of the infection episode.

An infection was considered symptomatic if the participant had reported a symptom onset date seven days pre or post the date of first positive PCR sample for each infection episode.

The proportion of participants with each infection was summarised by sociodemographic characteristics and vaccination status. Symptom type, number, duration, hospital attendance and time off work were compared by infection and vaccination status (SARS-CoV-2 and influenza only) using proportions.

The proportion of participants reporting ILI symptoms was calculated over time, by dividing the number of surveys where ILI was reported by the total number of surveys in each time period.

Sick leave rate was calculated by fortnight, using the number of days taken off work divided by the number of days participants contributed to the analysis period, per 100 days, with 95% confidence interval. Incidence rate ratio were calculated to estimate demographic factors associated with taking time off work, unadjusted and adjusted (vaccination status, age, occupational setting and co-morbidities).

For sick leave rate by vaccination status, participants contributed days to the vaccinated state (COVID-19 and influenza separately or simultaneously) if they had received the vaccine at least 14 days before the start of the fortnightly survey period. Participants contributed days to the unvaccinated state if they had not received the seasonal dose or had received the dose after the end of the survey period. For participants who received either vaccine within a survey period, or within the 14 days prior to the survey start date, this survey period did not count towards the rate analysis.

## RESULTS

A total of 7,774 participants consented to the SIREN Winter Pressures pilot sub-study from 28 November 2022 to 31 March 2023. Of these, 5,863 (75.4%) participants had both survey and testing data and were included in the analysis (Fig 1). Most participants were female (84.6%), of white ethnicity (91.4%), over 45-years of age (70.3%); nursing was the largest staff group (33.4%). Study participants were highly vaccinated, with 74.2% receiving both COVID-19 and the seasonal influenza vaccine (Table 1).

**Fig 1:**
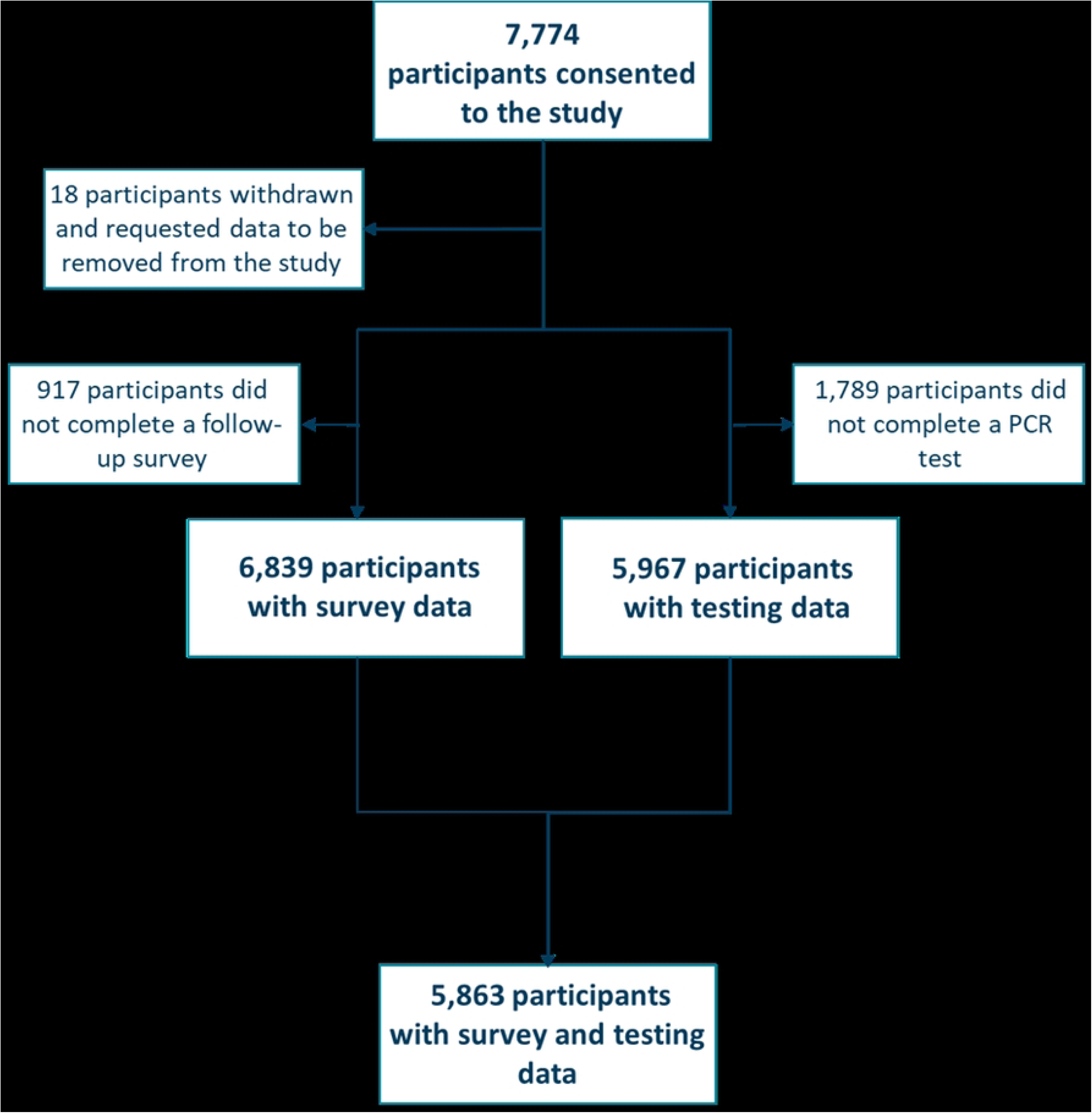
Flow diagram describing participants included in the SIREN Winter Pressures pilot sub-study.

**Table 1:** Demographics of participants included in the analysis, overall and by infection (SARS-CoV-2, Influenza and RSV), 28 November 2022 to 31 March 2023.

| Characteristic | All participants<br>n (%) | SARS-CoV-2<br>positive<br>N (%) | Influenza<br>positive<br>n (%) | RSV<br>positive<br>n (%) |
| --- | --- | --- | --- | --- |
| **Total** | **5,863** | **643** | **91** | **74** |
| **Age group** |  |  |  |  |
| Under 25 | 39 (0.7) | 1 (0.2) | 1 (1.1) | 3 (4.1) |
| 25 to 34 | 447 (7.6) | 56 (8.7) | 11 (12.1) | 2 (2.7) |
| 35 to 44 | 1,257 (21.4) | 151 (23.5) | 24 (26.4) | 13 (17.6) |
| 45 to 54 | 2,280 (38.9) | 244 (37.9) | 30 (33.0) | 36 (48.6) |
| 55 to 64 | 1,690 (28.8) | 173 (26.9) | 23 (25.3) | 20 (27.0) |
| Over 65 | 150 (2.6) | 18 (2.8) | 2 (2.2) | 0 (0.0) |
| **Gender** |  |  |  |  |
| Female | 4,960 (84.6) | 541 (84.1) | 67 (73.6) | 64 (86.5) |
| Male | 896 (15.3) | 101 (15.7) | 24 (26.4) | 9 (12.2) |
| Non-binary | 4 (0.1) | 1 (0.2) | 0 (0.0) | 1 (1.4) |
| Prefer not to say | 3 (0.1) | 0 (0.0) | 0 (0.0) | 0 (0.0) |
| **Ethnicity** |  |  |  |  |
| White | 5,359 (91.4) | 594 (92.4) | 85 (93.4) | 70 (94.6) |
| Asian | 272 (4.6) | 30 (4.7) | 4 (4.4) | 3 (4.1) |
| Black | 94 (1.6) | 5 (0.8) | 0 (0.0) | 0 (0.0) |
| Mixed Race | 70 (1.2) | 6 (0.9) | 1 (1.1) | 1 (1.4) |
| Other Ethnic Group | 52 (0.9) | 4 (0.6) | 1 (1.1) | 0 (0.0) |
| Prefer not to say | 16 (0.3) | 4 (0.6) | 0 (0.0) | 0 (0.0) |
| **Medical condition** |  |  |  |  |
| Chronic respiratory conditions | 738 (12.6) | 75 (11.7) | 13 (14.3) | 8 (10.8) |
| Chronic non-respiratory conditions | 679 (11.6) | 76 (11.8) | 8 (8.8) | 13 (17.6) |
| Immunosuppression | 128 (2.2) | 16 (2.5) | 2 (2.2) | 1 (1.4) |
| No medical condition | 4,318 (73.6) | 476 (74.0) | 68 (74.7) | 52 (70.3) |
| **Household** |  |  |  |  |
| Lives alone | 683 (11.6) | 82 (12.8) | 11 (12.1) | 9 (12.2) |
| Lives with adults | 3,010 (51.3) | 325 (50.5) | 42 (46.2) | 30 (40.5) |
| Lives with children | 2,170 (37.0) | 236 (36.7) | 38 (41.8) | 35 (47.3) |
| **Staff type** |  |  |  |  |
| Nursing | 1,957 (33.4) | 221 (34.4) | 22 (24.2) | 21 (28.4) |
| Administrative/Executive | 1,014 (17.3) | 95 (14.8) | 14 (15.4) | 11 (14.9) |
| Doctor | 670 (11.4) | 67 (10.4) | 12 (13.2) | 14 (18.9) |
| Healthcare Assistant | 360 (6.1) | 53 (8.2) | 3 (3.3) | 2 (2.7) |
| Healthcare Scientist | 279 (4.8) | 32 (5.0) | 7 (7.7) | 5 (6.8) |
| Student | 175 (3.0) | 25 (3.9) | 3 (3.3) | 1 (1.4) |
| Physiotherapist/Occupational Therapist/SALT | 228 (3.9) | 33 (5.1) | 10 (11.0) | 4 (5.4) |
| Midwife | 133 (2.3) | 12 (1.9) | 3 (3.3) | 1 (1.4) |
| Pharmacist | 129 (2.2) | 16 (2.5) | 3 (3.3) | 2 (2.7) |
| Estates/Porters/Security | 113 (1.9) | 8 (1.2) | 3 (3.3) | 1 (1.4) |
| Other | 805 (13.7) | 81 (12.6) | 11 (12.1) | 12 (16.2) |
| **Occupation setting** |  |  |  |  |
| Office | 1,333 (22.7) | 135 (21.0) | 26 (28.6) | 11 (14.9) |
| Outpatient | 1,213 (20.7) | 135 (21.0) | 19 (20.9) | 18 (24.3) |
| Inpatient Wards | 671 (11.4) | 89 (13.8) | 9 (9.9) | 13 (17.6) |
| Patient facing (non-clinical) | 273 (4.7) | 29 (4.5) | 2 (2.2) | 8 (10.8) |
| Intensive Care | 208 (3.5) | 30 (4.7) | 4 (4.4) | 4 (5.4) |
| Theatres | 152 (2.6) | 22 (3.4) | 2 (2.2) | 5 (6.8) |
| Ambulance/Emergency Department | 108 (1.8) | 12 (1.9) | 4 (4.4) | 2 (2.7) |
| Maternity/Labour Ward | 82 (1.4) | 7 (1.1) | 1 (1.1) | 1 (1.4) |
| Other | 1,823 (31.1) | 184 (28.6) | 24 (26.4) | 12 (16.2) |
| Patient contact |  |  |  |  |
| Yes | 4,841 (82.6) | 535 (83.2) | 75 (82.4) | 71 (95.9) |
| No | 1,022 (17.4) | 108 (16.8) | 16 (17.6) | 3 (4.1) |
| Vaccination status |  |  |  |  |
| Both | 4,349 (74.2) | 481 (74.8) | 62 (68.1) | 54 (73.0) |
| Influenza 2022/23 vaccine only | 499 (8.5) | 53 (8.2) | 8 (8.8) | 5 (6.8) |
| COVID-19 second booster<br>dose only | 354 (6.0) | 28 (4.4) | 10 (11.0) | 5 (6.8) |
| Neither | 661 (11.3) | 81 (12.6) | 11 (12.1) | 10 (13.5) |

*Note: Eight participants had multiple infections during the analysis period – four participants had a SARS-CoV-2 and an influenza infection, three had influenza and RSV, and one had SARS-CoV-2 and RSV.*

### SARS-CoV-2, influenza and RSV infection rates over winter 2022/23

All 5,863 participants were tested for SARS-CoV-2, 4,689 (80.0%) for influenza and 4,695 (80.0%) for RSV. Of these, 643/5,863 (11.0%) participants had a positive SARS-CoV-2 result, 93/4,689 (2.0%) had a positive influenza result and 74/4,695 (1.6%) had a positive RSV result.

PCR positivity rates for all three viruses peaked in early December 2022 (peak positive PCR per 100 tests: 4.6 (95% CI 3.5, 5.7) for SARS-CoV-2, 3.9 (95% CI 2.2, 5.6) influenza, 1.4 (95% CI 0.4, 2.4) RSV), with influenza and RSV decreasing to low levels after January 2023 (for influenza ≤0.2 and RSV ≤0.3 positives per 100 tests), whereas SARS-CoV-2 maintained rates around 2.5 per 100 tests until March 2023 (Fi 2).

**Fig 2:**
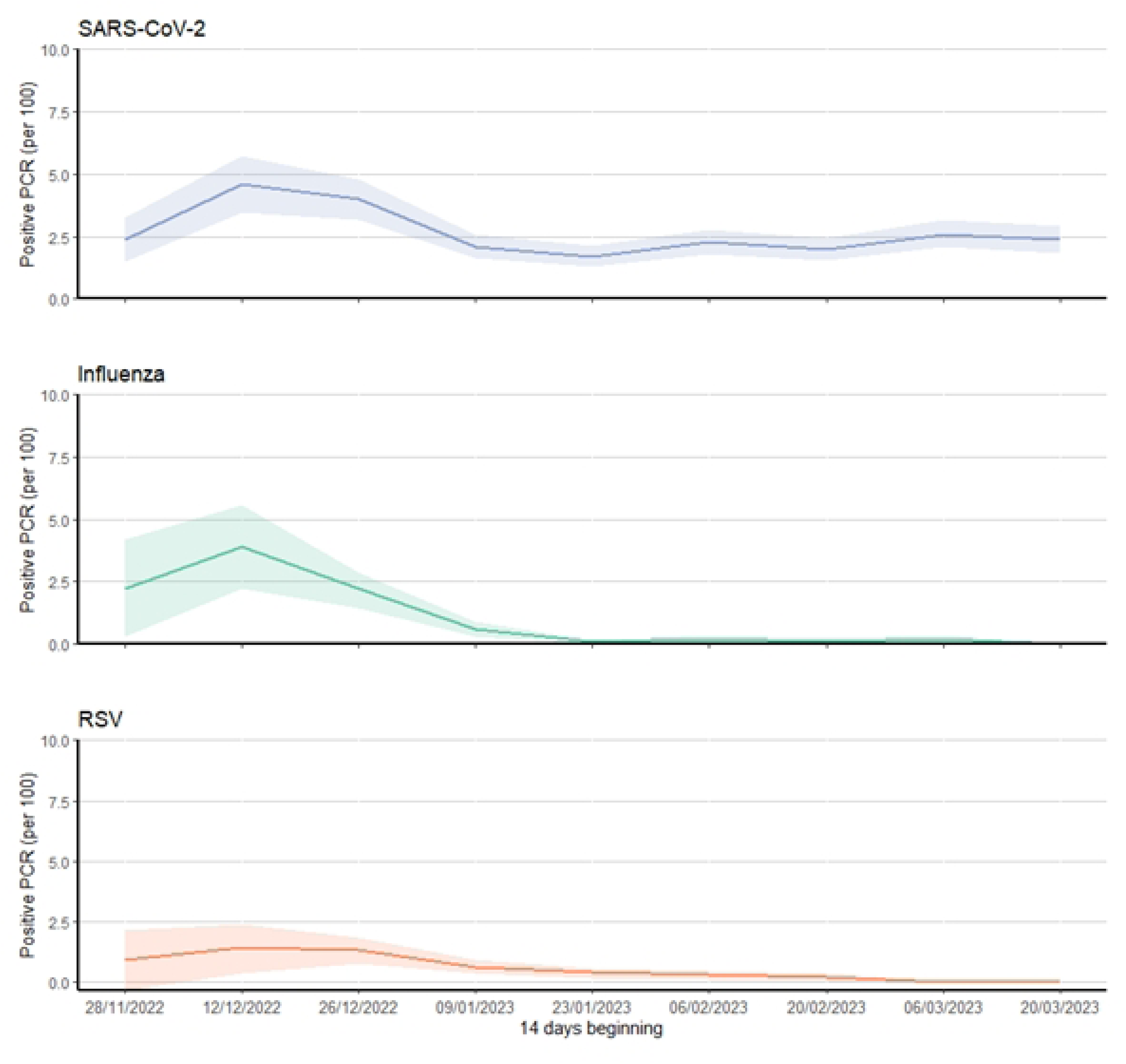
PCR positivity rate (per 100 tests) by fortnight and infection, 28 November 2022 to 31 March 2023. *Note: shaded areas represent 95% confidence intervals*

### Influenza-like illness symptoms over winter 2022/23

There were 4,003 (68.3%) participants who reported symptoms over the analysis period, with 1,054 (18.0%) reporting ILI symptoms. The proportion of participants reporting ILI symptoms by fortnight peaked in early December 2022 (peak fortnight was 7.5% (95% CI 6.8, 8.3) (368/4,893 surveys) (Fig 3).

**Fig 3:**
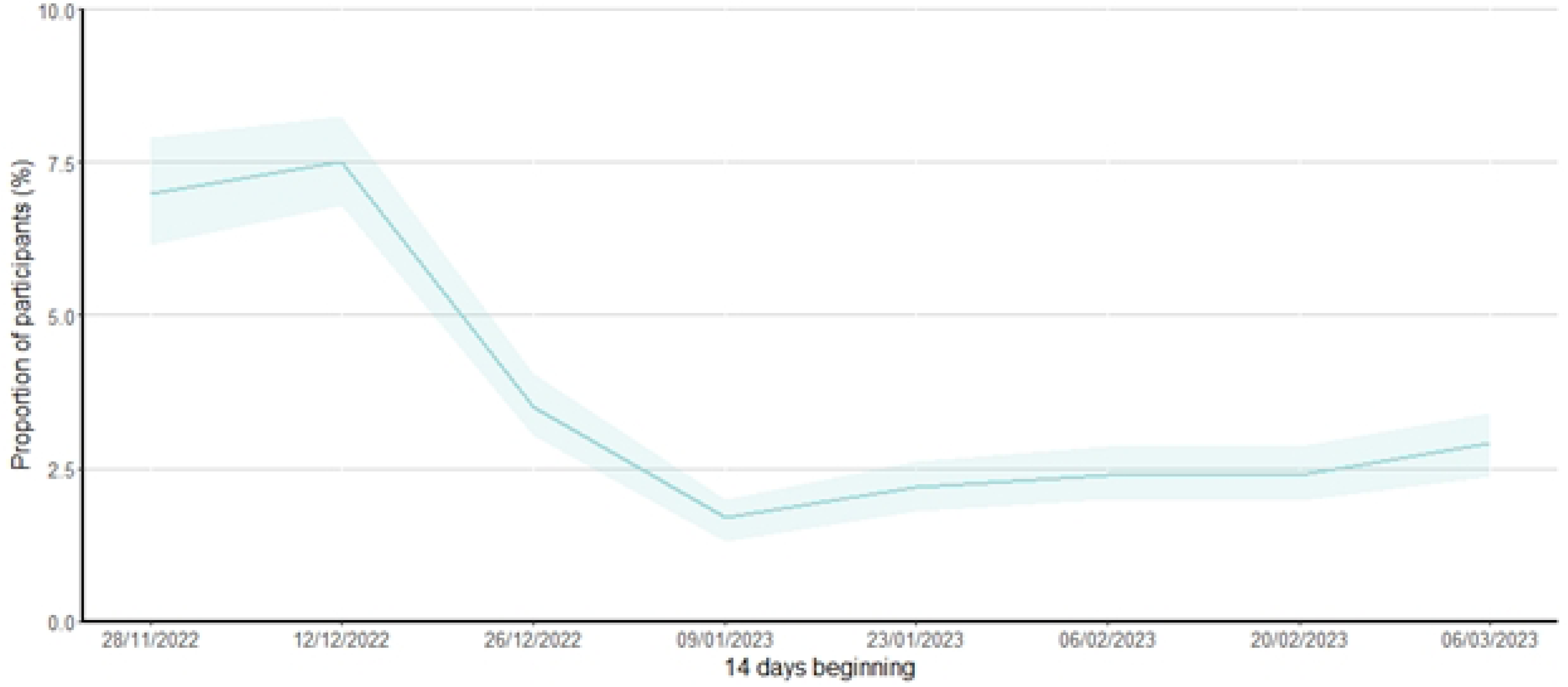
Proportion of participants reporting influenza-like illness symptoms by fortnight, 28 November 2022 to 31 March 2023. *Shaded areas represent 95% confidence intervals*

### Symptom profile by SARS-CoV-2, influenza and RSV

The proportion of asymptomatic infections were similar among the three viruses – influenza (45.6%), SARS-CoV-2 (41.7%) and RSV (37.5%). Symptom profiles were similar for all three viruses. Although, loss of sense of smell and taste were more common among those with an SARS-CoV-2 infection and fever more common with a influenza infection. Hospital attendance was low for all viruses (Table 2).

**Table 2:** Description of the symptom profile and severity of SARS-CoV-2, influenza and RSV PCR positive infections, 28 November 2022 to 31 March 2023.

|  | **SARS-CoV-2**<br>**n (%)** | **Influenza**<br>**n (%)** | **RSV**<br>**n (%)** | *p-value*<br>*(SARS-CoV-2*<br>*vs*<br>*influenza)* | *p-value*<br>*(SARS-CoV-2*<br>*vs RSV)* | *p-value*<br>*(RSV vs*<br>*influenza)* |
| --- | --- | --- | --- | --- | --- | --- |
| **Total** | **643** | **93** | **74** |  |  |  |
| **Symptoms** |  |  |  | *0.494* | *0.594* | *0.327* |
| Any symptoms | 346 (58.3) | 49 (54.4) | 40 (62.5) |  |  |  |
| Asymptomatic | 247 (41.7) | 41 (45.6) | 24 (37.5) |  |  |  |
| **Symptoms type** |  |  |  |  |  |  |
| Loss of smell | 67 (11.3) | 3 (3.3) | 4 (6.3) | *0.015* | *0.289* | *0.45* |
| Cough | 182 (30.7) | 33 (36.7) | 17 (26.6) | *0.274* | *0.568* | *0.223* |
| Diarrhoea | 37 (6.2) | 5 (5.6) | 2 (3.1) | *>0.999* | *0.414* | *0.7* |
| Loss of taste | 80 (13.5) | 3 (3.3) | 4 (6.3) | *0.005* | *0.116* | *0.45* |
| Fatigue | 160 (27.0) | 24 (26.7) | 13 (20.3) | *>0.999* | *0.297* | *0.445* |
| Fever | 105 (17.7) | 25 (27.8) | 9 (14.1) | *0.03* | *0.602* | *0.05* |
| Swollen glands | 45 (7.6) | 6 (6.7) | 1 (1.6) | *>0.999* | *0.074* | *0.24* |
| Headache | 228 (38.4) | 29 (32.2) | 24 (37.5) | *0.294* | *>0.999* | *0.606* |
| Muscle aches | 179 (30.2) | 24 (26.7) | 13 (20.3) | *0.538* | *0.112* | *0.445* |
| Runny nose | 258 (43.5) | 33 (36.7) | 28 (43.8) | *0.253* | *>0.999* | *0.406* |
| Shortness of breath | 87 (14.7) | 12 (13.3) | 13 (20.3) | *0.873* | *0.27* | *0.273* |
| Sore throat | 231 (39.0) | 39 (43.3) | 29 (45.3) | *0.488* | *0.348* | *0.87* |
| Vomiting | 38 (6.4) | 8 (8.9) | 5 (7.8) | *0.368* | *0.598* | *>0.999* |
| **Duration of symptoms** |  |  |  | *0.09* | *0.405* | *0.729* |
| 1-3 | 43 (12.6) | 1 (2.0) | 2 (5.0) |  |  |  |
| 4-6 | 79 (23.2) | 10 (20.4) | 8 (20.0) |  |  |  |
| 7-14 | 54 (15.9) | 8 (16.3) | 9 (22.5) |  |  |  |
| >14 | 164 (48.2) | 30 (61.2) | 21 (52.5) |  |  |  |
| **Sick leave** |  |  |  | *0.002* | *0.868* | *0.049* |
| Taken | 180 (52.2) | 14 (28.6) | 20 (50.0) |  |  |  |
| Not taken | 165 (47.8) | 35 (71.4) | 20 (50.0) |  |  |  |
| **Number of sick leave taken (days)** |  |  |  | *0.015* | *0.484* | *0.079* |
| 1-3 | 67 (37.2) | 9 (64.3) | 10 (50.0) |  |  |  |
| 4-6 | 56 (31.1) | 5 (35.7) | 4 (20.0) |  |  |  |
| 7-14 | 57 (31.7) | 0 (0.0) | 6 (30.0) |  |  |  |
| **Hospital attendance** |  |  |  | *0.358* | *0.095* | *0.654* |
| Yes | 8 (2.3) | 2 (4.1) | 3 (7.5) |  |  |  |
| No | 338 (97.7) | 47 (95.9) | 37 (92.5) |  |  |  |

### Time off work due to being symptomatic over winter 2022/23

Of participants testing positive for SARS-CoV-2 and RSV, ≥50% of participants reported taking time off work for being symptomatic (52.2% and 50.0%, respectively), compared to 28.6% for influenza infections (Table 2). The median number of days taken off work for a SARS-CoV-2 infection was 5 (IQR: 2-7); 3 days (IQR: 2.25-5) for influenza and 4 days (IQR: 2-7) for RSV.

Over the analysis period, a total of 1,368/5,863 (23.3%) participants reported taking time off work due to being symptomatic; taking a median of 4 days off over the analysis period (range: 1-59 days). Of participants reporting ILI symptoms, 616/1,054 (58.4%) reported taking time off work; taking a median of 5 days; range: 1-59 days.

The 5,863 participants included contributed to a total of 507,388 follow-up days during the winter period. The rate of sick leave over this period was 1.48 days per 100 days of follow-up, with the peak seen in early December 2022 (2.4 days per 100 days) (Fig 4).

**Fig 4:**
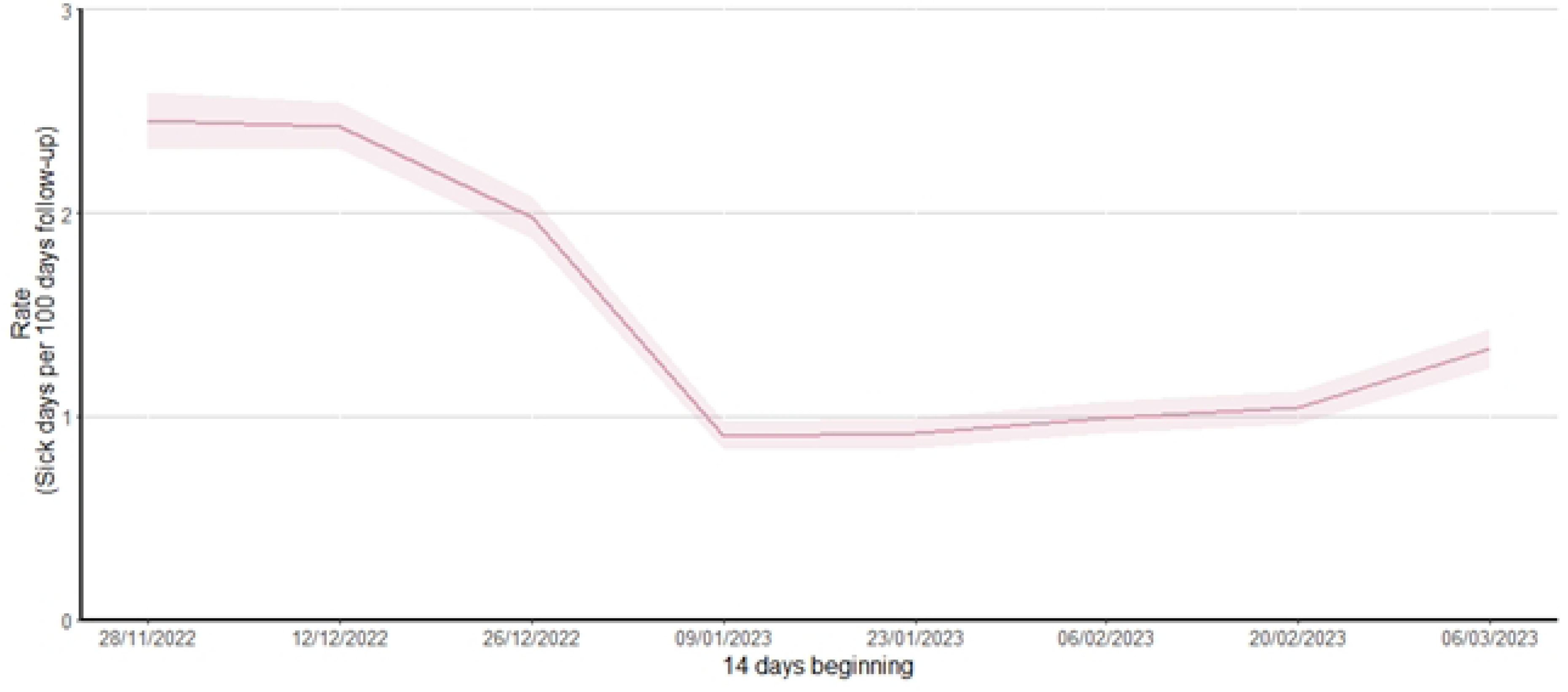
Rate of sick leave taken (per 100 days) by fortnight, 28 November 2022 to 31 March 2023. *Note: Sick leave rate was calculated as number of sick leave days taken divided by the number of days participants contributed to the analysis period, per 100 days; Shaded areas represent 95% confidence intervals*

### Factors associated with taking time off work

Participants who took time off work due to being symptomatic between November 2022 and March 2023 appeared to differ by occupational setting, long-term medical conditions and vaccination status (Table 3).

**Table 3:**
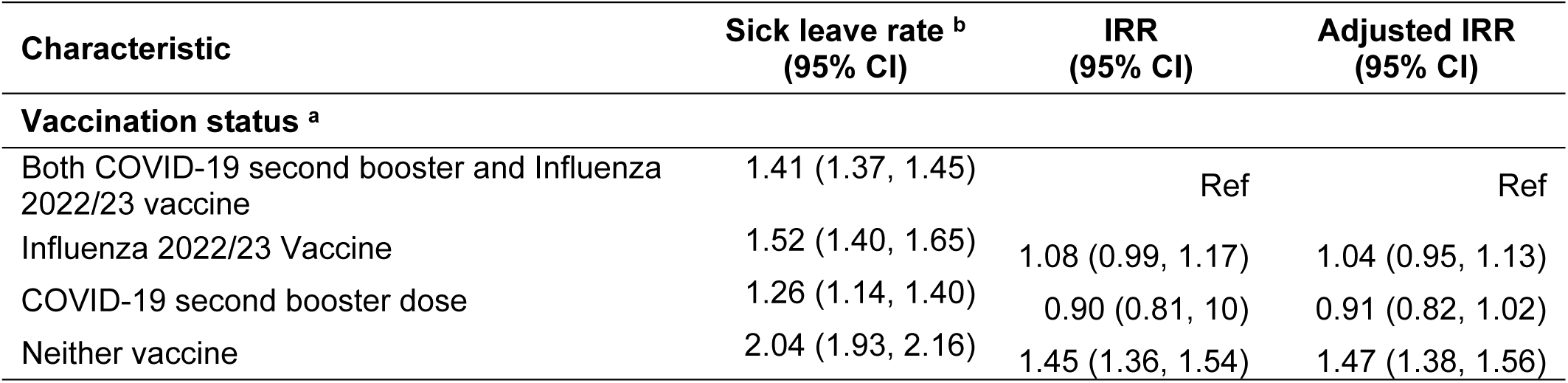
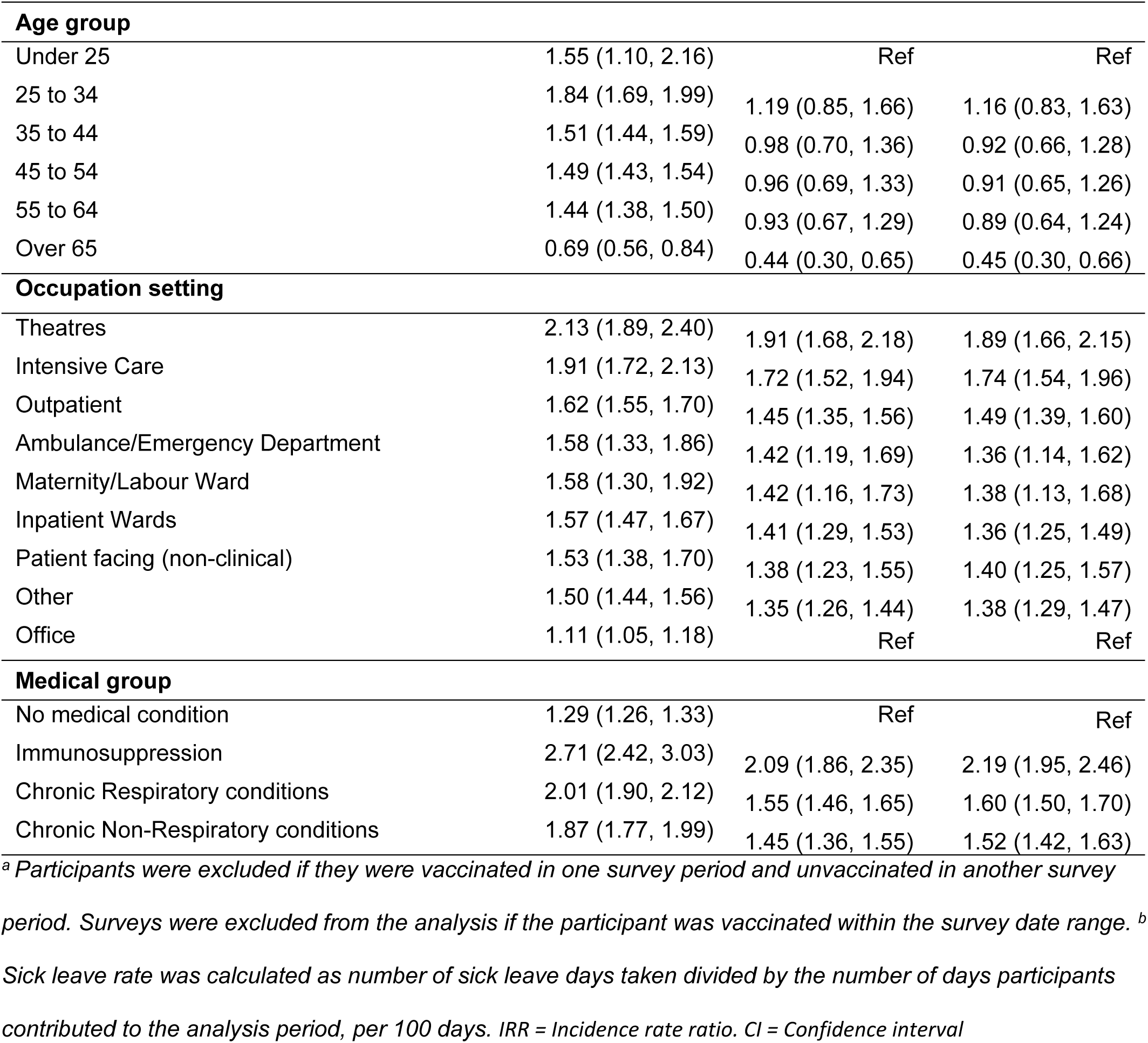
Factors associated with time off work due to being symptomatic, 28 November 2022 to 31 March 2023.

Participants in office-based roles took less time off work (1.11 days per 100 days) when compared to clinical roles such as theatres and intensive care units (2.13 and 1.91 days per 100 days, respectively. Estimated from adjusted Incidence Rate Ratios (IRR) also show this; theatres (1.89; 95% CI 1.66, 2.15) and intensive care units (1.74; 95% CI 1.54, 1.96) verses office-based participants. Participants with immunosuppressive, chronic respiratory and chronic non-respiratory conditions took more time off work than participants with no reported comorbidities (who took off 1.29 days per 100 days): immunosuppressive conditions 2.71 days per 100, adjusted IRR 2.19 (95% CI 1.95, 12.46); chronic respiratory 2.01/100 days, adjusted IRR 1.60 (95% CI 1.50, 1.70); and chronic non-respiratory conditions 1.87/100 days, adjusted IRR 1.52 (95% CI 1.42, 1.63).

Those who were vaccinated against both influenza and COVID-19 took less time off work (1.41 days per 100 days) than those who did not receive either vaccine (2.04 days per 100 days) (Table 3). After controlling for age, occupational setting and co-morbidities, we estimated that those who received neither vaccine had a 47% higher rate of sick leave than those vaccinated for both (adjusted IRR 1.47 (95% CI 1.38, 1.56)).

## DISCUSSION

Results from the 2022/23 SIREN Winter Pressures pilot study demonstrate the impact of respiratory illness on the NHS workforce. Rates of SARS-CoV-2, influenza and RSV, and symptoms, all peaked in early December, resulting in increased levels of sickness absence over this time period.

The PCR positivity trends over winter 2022/23 in our cohort of HCW is consistent with national surveillance data (17) demonstrating the potential utility of conducting surveillance in this population to determine the prevalence and impact of respiratory viral infections in the working age population (20). Our detection of RSV infections in this population, which is rarely tested for this virus, suggests that RSV also contributes to winter pressures. The finding that symptom profiles were similar across the three viruses is consistent with existing published literature (21). Across all three viruses, a substantial proportion of infections were asymptomatic but could potentially contribute to transmission, in particular, influenza with over two-thirds of infections asymptomatic (22). This highlights the potential risk associated with nosocomial infections in healthcare workers, though we do not have strong evidence how transmissible asymptomatic infections is. The impact of respiratory illness on time off work in our cohort of HCW over winter 2022/23 was considerable, with 23% of participants reporting time off work due to being symptomatic, and all three viruses contributing to this. Sick leave rates varied by demographics and vaccination status, with lower estimated rates among those who received the second COVID-19 booster and seasonal influenza vaccine. Given the impact that HCW time off work could have on healthcare resilience over winter, further research into associated factors, including behaviours and attitudes, is important.

A key limitation of this pilot study was the coverage and timing of multiplex PCR roll-out. There were fewer participants with PCR results for influenza and RSV than those with a SARS-CoV-2 test result. In this pilot, multiplex testing was introduced late in the winter season, with testing data only available from late November 2022, due to delays switching PCR platforms across NHS laboratories (using a decentralised study testing model), and the timing of establishing a new centralised postal PCR pathway. This delay was compounded by an unusually early influenza season in 2022/23 (17). Consequently, our surveillance of these viruses over winter 2022/23 may have missed some infections within our cohort, particularly influenza and RSV.

This pilot sub-study conducted during winter 2022/23 has demonstrated the adaptability and applicability of findings from the SIREN HCW cohort. Results from the study show the benefit of regular multiplex testing across NHS Trusts to understand the interplay and impact of seasonal viruses on workforce planning, patient care and healthcare resilience in the NHS during the Winter. We have demonstrated that all three viruses contribute to staff illness and time off work over winter, and that seasonal flu and COVID-19 vaccines were associated with lower sick leave rates. Future studies should consider using a centralised multiplex testing pathway to improve surveillance of respiratory infections and ensure the timing of testing is optimised.

## Data Availability

Anonymised data will be made available for secondary analysis to trusted researchers upon reasonable request.

## ACKNOWLEDGEMENTS

Our thanks go to the participants in the SIREN study. We would also like to thank the help and support offered by the Berkshire Research Ethics Committee and the HDR-UK for winter pressures funding and to UKHSA for core SIREN study funding. Thanks also to all research team and site staff not mentioned above.

